# Association of Select Psychiatric Disorders with Incident Brain Aneurysm and Subarachnoid Hemorrhage among Veterans

**DOI:** 10.1101/2023.02.26.23286479

**Authors:** Daniel L Cooke, Hui Shen, Madhavi Duvvuri, Daniel Thompson, Thomas Neylan, William Wolfe, Steven Hetts, Bruce Ovbiagele, Mary Whooley, Beth Cohen

**Affiliations:** University of California San Francisco, Dept. of Radiology and Biomedical Imaging; University of California San Francisco, School of Medicine, San Francisco Veterans Affairs Medical Center; University of California San Francisco, Dept. of Psychiatry, San Francisco Veterans Affairs Medical Center; University of California San Francisco, Dept. of Neurology, San Francisco Veterans Affairs Medical Center; University of California San Francisco, Dept of Medicine, San Francisco Veterans Affairs Medical Center

## Abstract

**Background:** Brain aneurysms represent a significant cause of hemorrhagic stroke. Prior research has demonstrated links between stress and stroke, including brain aneurysms. We aimed to determine relationships between select psychiatric disorders and aneurysms and aneurysmal SAH.

**Methods:** We performed retrospective, case-control study of a National Veterans Affairs population with two experimental groups (aneurysm-only and aneurysmal SAH) and 10-fold controls per group matched by age, date, and clinical data source. The studied the presence of 4 psychiatric disorders: Posttraumatic stress disorder (PTSD), major depressive disorder (MDD), generalized anxiety disorder (GAD), and other mood disorders. Our main outcomes Unadjusted and multivariable adjusted ORs of PTSD, MDD, GAD, and mood disorders within aneurysm-only and aSAH groups.

**Results:** In 6,320,789 US Veterans who were enrolled for at least 5 years in Medicare and/or the Veterans Health Administration, we identified 35,094 cases of aneurysm without SAH and 5749 cases of aneurysm with SAH between 1/2005 and 12/2019. In analyses adjusted for sex, hypertension, and tobacco use, patients with aneurysm were more likely than matched controls to have a history of PTSD (OR 1.48), MDD (OR 1.33), GAD (OR 1.26), and other mood disorders (OR 1.34) (all p values <0.0001). Similarly, patients with aSAH were more likely than controls to have a history of PTSD (OR 1.35), MDD (OR 1.38), GAD (OR 1.18), and other mood disorders (OR 1.30) (all p values <0.0001).

**Conclusions:** The study, the largest of its kind, further suggests links between psychiatric disorders and stroke. This is important as patients with aneurysms are not routinely screened for such psychiatric risk factors. Additional research on this topic could lead to novel strategies to improve stroke prevention.

## Introduction

Aneurysmal subarachnoid hemorrhage (aSAH) carries significant morbidity and is fatal in approximately 30% of cases ^1,2^. Brain aneurysms are relatively common with an estimated prevalence of 1%–3% depending on the adult population ^3,4^. Aneurysm size and genetic factors have been shown to play a role in rupture while several other factors, including smoking and hypertension, also influence risk and have informed models of stroke. Identification and management of such underlying modifiable risk factors and exposures have led to a global decline of aSAH ^5^. Within this same 20-year interval (1993 – 2015), no significant change in the annual incidence of aSAH has occurred within the US ^6^ despite an increase in the elective treatment of aneurysms overall. As such, the neurovascular community continues to search for other patient characteristics, such as female sex, biomarkers (e.g., MR-based aneurysmal wall enhancement) and modifiable risk factors (e.g., drug use), to more reliably differentiate at-risk subgroups as effectively as aneurysmal size.

The role of psychosocial stress in stroke is well established, and neuropsychiatric disorders, including major depressive disorder and schizophrenia, are considered significant risk factors for adverse clinical events ^7-9^. Mechanisms through which mental health disturbances increase stroke risk have not, however, been fully elucidated. Evidence indicates roles for refractory hypertension, inflammation, autonomic nervous system disturbance, and/or endocrine dysfunction ^7,10^,^11^. Despite sharing pathophysiological mechanisms (e.g., atherosclerosis) and risk factors (e.g., hypertension) with myocardial infarction and ischemic stroke, a connection between aneurysms, aSAH and psychiatric disorders and environmental stressors has yet to be well-established. Significant financial and domestic stress has been associated with aneurysm presence and rupture, although these findings need validation in larger samples ^12^. Recent work has also demonstrated a possible interaction between PTSD, among other psychiatric disorders, and hemorrhagic stroke in young Veterans ^13^. After adjusting for demographics, health behaviors, traditional cardiovascular risk factors, and psychiatric comorbidities, PTSD proved non-influential in risk modeling. However, this study had a relatively small percentage of subarachnoid hemorrhage patients and did not address aneurysms directly.

Given the need for better risk stratification of patients with unruptured aneurysms and an incomplete understanding of the interaction between aneurysms, aSAH, and psychiatric disorders, we performed a retrospective case-control study of a large United States Veteran population to examine psychological conditions as predictors of aneurysms and aSAH.

## Methods

### Sources of Data

We utilized the STROBE checklist for case-control studies for manuscript development^14^ and adhered to observation case-control study guidelines. We used data from the Veterans Health Administration (VA) Corporate Data Warehouse (CDW). The CDW contains information on VA inpatient and outpatient visits and associated clinical diagnoses and procedures, information on non-VA visits reimbursed by VA, vital signs, VA pharmacy records, laboratory data, and other patient-level variables. We also used Medicare data from the Centers for Medicare and Medicaid Services (CMS) for all Veterans enrolled in or eligible for VHA healthcare and linked these data to VA data.

### Participants

We examined records from 2000 to 2019 to identify patients eligible for VA care who were enrolled in VHA or Medicare for at least 60 months and had at least 1 VA visit during that time period (Figure 1). Patients enrolled in Medicare Part C at any time during the study period were excluded from the analysis because we only had data available for Medicare Parts A and B. We used ICD and CPT codes (Supplemental Table 1) to identify two case groups: 1. Brain aneurysm only and 2. Brain aneurysm plus non-traumatic SAH. Patients needed 1 ICD code from inpatient visits, 1 procedure code, or 2 outpatient codes qualify. For aneurysm we used codes ICD-9 437.3, ICD-10 I67.1, and CPT codes 61697, 61698, 61700, 61702, 61624. For non-traumatic SAH we used codes ICD-9 430 and ICD-10 I60. We then randomly selected 10 controls per case from remaining patients who did not have any aneurysm or SAH codes.

**Fig. 1:**
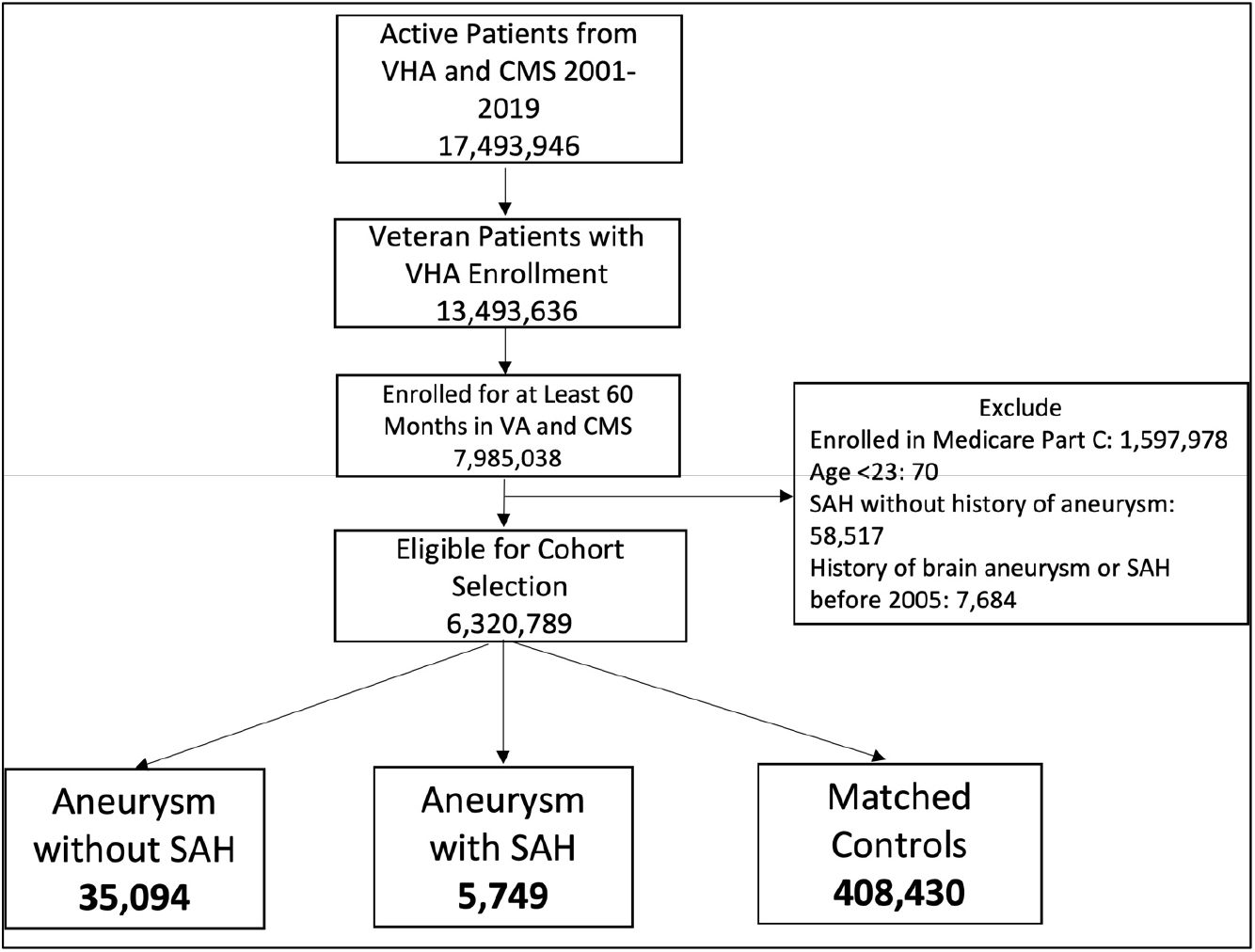
Flow chart of patient selection methods.

Cases and controls were matched by age, date of diagnosis (case diagnosis matched to a clinical visit date for controls), and site of index visit (VA vs. CMS). After applying selection criteria, 6,320,789 patients were eligible for inclusion. The case and control selection and matching process yielded 35,094 aneurysm-only and 5749 aSAH patients and 10x matched control patients for each of these groups (Figure 1).

This study was approved by the University of California, San Francisco Human Research Protection Program and the San Francisco Veterans Affairs Healthcare System Research and Development Committee. As the study involved secondary analysis of healthcare records, no participant contact was involved, and informed consent was not required.

### Variables

We evaluated the 5-year period prior to the index date to establish diagnoses of several mental health conditions using ICD codes: PTSD, major depressive disorder (MDD), generalized anxiety disorder (GAD), and other mood disorders (Appendix Table 1). We collected additional demographic information (age and gender) and used the same prior 5-year period to assess diagnoses of common comorbidities and stroke risk factors (hypertension, diabetes, and tobacco use).

### Statistical Analyses

Differences in patient characteristics between cases and controls were compared using the Chi-square test for categorical variables and Wilcoxon rank-sum test for continuous variables. Univariable logistic regression models were constructed for each mental health predictor with the outcome of brain aneurysm. Analogous models were constructed with the outcome of aSAH. Subsequent models included all psychiatric factors simultaneously to examine the independent associations of the mental health predictors with the outcomes. Multivariable models adjusted for sex, hypertension, diabetes, and tobacco use. All analyses were performed with SAS Enterprise Guide version 8.3 (SAS Institute, Cary, NC).

## Results

Patients with Aneurysm-only were older than those with aSAH (70.8 years vs 68.1 years). Female sex, hypertension, and tobacco use were more common in both aneurysmal groups relative to controls (Tables 1 and 2, p-value < 0.0001).

**Table 1:** Characteristics of participants with <u>aneurysm only</u> versus controls matched for age, date and site of diagnosis (VA or CMS)

|  |  |  |  |
| --- | --- | --- | --- |
| **Variable** | **Aneurysm Only (N=35094)** | **VA/CMS patients Age and Site Matched to Aneurysm only Patients (N=350940)** | **p-value** |
| **Age at first aneurysm diagnosis date/matched clinic visit date: Mean +- STD** | 70.76+-12.56 | 70.76+-12.56 | 1.00** |
| **Female** | 2398 (6.8%) | 14838 (4.2%) | <0.0001 |
| **Hypertension** | 27915 (79.5%) | 254128 (72.4%) | <0.0001 |
| **Diabetes mellitus** | 12563 (35.8%) | 119258 (34.0%) | <0.0001 |
| **Tobacco Use Disorder** | 22892 (65.2%) | 205659 (58.6%) | <0.0001 |
| **Generalized Anxiety Disorder** | 4420 (12.6%) | 33607 (9.6%) | <0.0001 |
| **Depression** | 9720 (27.7%) | 74385 (21.2%) | <0.0001 |
| **Other mood disorders** | 1713 (4.9%) | 11868 (3.4%) | <0.0001 |
| **Post-traumatic stress disorder** | 2104 (6.0%) | 13520 (3.9%) | <0.0001 |
| P-values were calculated from Chi-square test for category variables and ANOVA for continuous variables unless otherwise marked<br>* P-values were calculated from Fisher's Exact test<br>** Variable was rank transformed |  |  |  |

**Table 2:** Characteristics of participants with <u>aneurysm and SAH</u> vs. controls matched for age, date and site of diagnosis (VA or CMS)

| **Variable** | **Both Aneurysm and SAH (N=5749)** | **VA/CMS patients Age and Site Matched to SAH Patients (N=57490)** | **p-value** |
| --- | --- | --- | --- |
| **Age at first aneurysm diagnosis date/matched clinic visit date: Mean +- STD** | 68.10+-12.83 | 68.10+-12.83 | 1.00** |
| **Female** | 475 (8.3%) | 2918 (5.1%) | <0.0001 |
| **Hypertension** | 4501 (78.3%) | 39897 (69.4%) | <0.0001 |
| **Diabetes mellitus** | 1812 (31.5%) | 18760 (32.6%) | 0.09 |
| **Tobacco Use Disorder** | 3997 (69.5%) | 33926 (59.0%) | <0.0001 |
| **Generalized Anxiety Disorder** | 730 (12.7%) | 5758 (10.0%) | <0.0001 |
| **Depression** | 1730 (30.1%) | 12676 (22.0%) | <0.0001 |
| **Other mood disorders** | 327 (5.7%) | 2237 (3.9%) | <0.0001 |
| **Post-traumatic stress disorder** | 354 (6.2%) | 2422 (4.2%) | <0.0001 |
| P-values were calculated from Chi-square test for category variables and ANOVA for continuous variables unless otherwise marked<br>* P-values were calculated from Fisher's Exact test<br>** Variable was rank transformed |  |  |  |

In univariate analysis, all psychiatric disorders were more common in patients with Aneurysm-only (Table 1) and aSAH (Table 2) than in controls. In analyses adjusted for hypertension, tobacco use, and sex, patients with Aneurysm-only were more likely than age- and site-matched controls to have a history of PTSD (OR 1.48), MDD (OR 1.33), GAD (OR 1.26), and other mood disorders (OR 1.34) (Table 3; all p values < 0.0001). Similarly, patients with aSAH were more likely than age- and site-matched controls to have a history of PTSD (OR 1.35), MDD (OR 1.38), GAD (OR 1.18), and other mood disorders (OR 1.30) (Table 4). In secondary multivariable models incorporating all psychiatric disorders simultaneously, all psychiatric variables remained independently associated with Aneurysm-only (Supplemental Table 2), but only MDD and other mood disorders remained independently associated with aSAH (Supplemental Table 3).

**Table 3:** Unadjusted and Adjusted Associations of Mental Health Conditions with Brain Aneurysm Only

|  |  |  |  |  |
| --- | --- | --- | --- | --- |
|  | **Unadjusted OR (95% CI)** | **p-value** | **Adjusted\* OR (95% CI)** | **p-value** |
| **PTSD** | 1.59<br>(1.52-1.67) | <0.0001 | 1.48<br>(1.41-1.56) | <0.0001 |
| **Depression** | 1.42<br>(1.39-1.46) | <0.0001 | 1.33<br>(1.30-1.36) | <0.0001 |
| **Generalized Anxiety Disorder** | 1.36<br>(1.32-1.41) | <0.0001 | 1.26<br>(1.22-1.30) | <0.0001 |
| **Other Mood Disorders** | 1.47<br>(1.39-1.54) | <0.0001 | 1.34<br>(1.27-1.41) | <0.0001 |
| *All models adjusted for sex, hypertension, diabetes, and tobacco use |  |  |  |  |

**Table 4:** Unadjusted and Adjusted Associations of Mental Health Conditions Aneurysm with Subarachnoid Hemorrhage

|  |  |  |  |  |
| --- | --- | --- | --- | --- |
|  | **Unadjusted OR (95% CI)** | **p-value** | **Adjusted\* OR (95% CI)** | **p-value** |
| **PTSD** | 1.49<br>(1.33-1.67) | <0.0001 | 1.35<br>(1.20-1.52) | <0.0001 |
| **Depression** | 1.52<br>(1.43-1.62) | <0.0001 | 1.38<br>(1.30-1.47) | <0.0001 |
| **Generalized Anxiety Disorder** | 1.31<br>(1.20-1.42) | <0.0001 | 1.18<br>(1.09-1.28) | <0.0001 |
| **Other Mood Disorders** | 1.49<br>(1.32-1.68) | <0.0001 | 1.30<br>(1.15-1.47) | <0.0001 |
| *All models adjusted for sex, hypertension, diabetes, and tobacco use |  |  |  |  |

## Discussion

The study represents the largest examination of the intersection of psychiatric disorders and brain aneurysms and aSAH. The results expand upon prior work implicating hypertension, smoking, and female sex as aneurysm risk factors ^15^. In multivariable modeling, PTSD, MDD, GAD, and other mood disorders were all significantly associated with aneurysms. Associations with psychiatric disorders were generally less pronounced within the aSAH group relative to the aneurysm-only group. This latter result may reflect under-recording of psychiatric disorders in non-VA healthcare facilities as most ruptured aneurysms are managed outside the VA system. Post-traumatic stress disorder and MDD proved the strongest predictors of aneurysm or aSAH, respectively. This is important to the stroke community as most patients with aneurysms are not screened for PTSD or MDD signs and symptoms as part of aneurysmal workup. Further, therapies, pharmacological or behavioral, dedicated to the treatment of such disorders, are not typically considered a stroke risk reduction strategy.

As noted, several studies have suggested a role for psychiatric disturbance in stroke ^16,17^ one of which found an association of aneurysms with hemorrhagic stroke, though it was not specific to SAH ^12^. In comparison to the study by Gaffey and colleagues, the current study included a significantly older population (~70 years vs. 30 years) with a greater prevalence of hypertension (~80% vs. 13%). Such age discrepancies may have contributed to differences in the fraction of psychiatric diagnoses, with GAD (~12% vs. 16%) and PTSD (~6% vs. 28%) being more common relative to the current work and MDD (~30 vs. 10%) being less so. Importantly, of hemorrhagic stroke subtypes, only 104 patients (25%) were SAH in nature and thus conclusions from this prior study regarding aneurysmal pathophysiology were more limited.

### Mechanisms

Several factors have been proposed as mechanisms through which mental health disorders could affect aneurysm growth and rupture, including hypertension, inflammation, oxidative stress, and immunoregulatory dysfunction. These mechanisms, whether independently or synergistically, can have a negative effect on the molecular-cellular environment of aneurysms, eventually leading to rupture. Hypertension, a consistent risk factor for aneurysms may result from stress-induced hypothalamic-pituitary axis dysfunction. This increases the secretion of glucocorticoids and circulating catecholamines, which over time leads to vascular smooth muscle cell modulation and endothelial damage^18^.

Neuropsychiatric disorders have shown association with elevated inflammation, as demonstrated by increases in C-reactive protein, interleukins, and fibrinogen ^18,19^. Aneurysmal pathophysiological modeling suggests following an initial insult to the endothelium due to hemodynamic wall stress, upregulation of the inflammatory pathways may initiate a feed-forward injury cycle. The chronic translocation of leukocytes through the endothelium can accelerate degradation of the vessel wall, making the aneurysms more prone to rupture. Levels of proteins such as vascular adhesion molecule 1 and von Willebrand factor, which are important in the leukocyte translocation pathway, have been shown to be higher in individuals with depression^20^. Additionally, TNF-a, a cytokine implicated in aneurysm pathogenesis^21^, has been shown to be elevated in patients with PTSD and MDD. TNF-a upregulates signaling pathways that lead to apoptosis and cellular degradation, such as the expression of calcium channels, Toll-like receptors, and matrix metalloproteinases. TNF-a and interleukin 1 signal downstream pro-inflammatory cytokines and interleukins while MMPs increase vascular permeability, leading to hemorrhage^22^. Such neurovascular immunological dysfunction may be mitigated in part through flow- and pressure-dependent triggers, though the upstream genetics and may offer opportunities to modify risk either through medical and/or lifestyle changes.

Beyond these inflammatory and immunoregulatory alterations, environmental, occupational and psychosocial stress can increase reactive oxygen species (ROS) and cause vascular dysfunction^23^. In mouse models, cigarette smoke exposure is increases ROS and NOX1 which precipitates upregulation of proinflammatory/matrix remodeling genes, leading to vascular smooth muscle cell modulation and aneurysm formation and rupture^24^. Increased oxidative stress and ROS production also affects the renin-angiotensin system and glucocorticoid pathways with downstream effects on endothelial cell function^25^. Moreover, the upregulation of free radicals, superoxide dismutase and glutathione in the setting of occupational and domestic stress is also associated with MDD^26^. Though no studies have specifically addressed these mechanisms in patients with aneurysms, studies of patients with cardiovascular risk factors have demonstrated stress reduction can reduce oxidative stress and improve markers of endothelial function^27^.

Lastly, patients with psychiatric disorders may disproportionately use and/or abuse substances modifying stroke risk. Chronic alcohol use has been implicated as a risk factor for hypertension and stroke^28^ and intensity of alcohol use has also been shown to demonstrate a dose-dependent relationship to aneurysm rupture^29^. Heroin and methamphetamine use have also been implicated in case series of aSAH patients^30,31^. It is possible that pharmacological interventions for psychiatric disorders may also affect stroke risk. There is controversy about how selective serotonin reuptake inhibitors may affect stroke with or without other drug modifiers^13,32^. More comprehensive, prospective studies will be needed to elucidate these potentially confounding relationships.

### Limitations

As this is an observational study, we cannot prove that psychiatric disorders caused aneurysms or aSAH. We relied on medical records to establish diagnoses of predictors and outcomes, and this could be subject to misclassification bias. Our findings may also be influenced by ascertainment bias if patients diagnosed with mental health disorders were more likely to seek care (e.g., undergo brain imaging) and have aneurysms diagnosed. However, aneurysm rupture would likely lead to symptoms that prompted treatment-seeking independent of prior mental health and treatment status. It is encouraging that established risk factors such as hypertension, female sex, and tobacco use were also associated with aneurysmal diagnoses suggesting the results might more accurately reflect reality. Finally, though we attempted to match or control for important factors that are associated with mental health disorders and could impact aneurysm and aSAH risk, residual confounding remains a concern.

## Data Availability

The data are owned by the Department of Veterans Affairs (VA) and cannot be shared publicly. Information about access to VA data can be found at: https://www.data.va.gov/ and https://www.virec.research.va.gov/. The authors do not have ownership of the data or the authority to execute a data use agreement. Interested researchers may contact [Name and email address of corresponding author], San Francisco VA Health Care System, for guidance on how to request individual data use agreements with the Department of Veterans Affairs.

## Acknowledgements

The authors wish to recognize the veterans themselves as well as the clinical and research staff at the SFVAMC.

## Supplementary Materials

Tables S1-S3

